# Glycovariants of CA-125 in the early detection of ovarian cancer

**DOI:** 10.64898/2026.09.11.26362728

**Authors:** Andrew J. Vickers, Olle Melander, Kara Long Roche, Katri Kuningas, Marjut Helle, Kaisa Huhtinen, Leena Kokko, Anders Dahlin, Hans Lilja

## Abstract

**Importance:** Two randomized trials have shown that screening with CA-125 is not of value for reducing ovarian cancer mortality. This appears to be because changes in CA-125 occur too late in the disease process to allow for surgical cure of early-stage disease.

**Objective:** To determine whether glycovariants of CA-125 could detect ovarian cancer years before clinical diagnosis.

**Design:** Case-control study of cryopreserved blood samples and data from the population-based Malmö Diet and Cancer cohort.

**Setting:** Population-based study in Sweden.

**Participants:** Serum was available for 17,297 women aged 44 – 73 collected at baseline in 1991-1996 and from 2,168 women re-sampled in 2007-2012. Participants’ records were linked to the Swedish cancer registry to identify 82 women with incident ovarian cancer diagnosed up to 10 years after blood draw. Cases were matched 1:3 with controls by age and venipuncture date.

**Exposure:** Serum was measured for CA-125 glycovariants Sialyl-Thomsen-nouveau (STn) and Macrophage-Galactose-Lectin (MGL) using GLYVAR^®^ Ovarian I and II assays (Uniogen), conventional CA-125 (CanAg CA125 EIA, Fujirebio) and HE4 EIA (Fujirebio).

**Main outcome measure:** Ovarian cancer.

**Results:** The CA-125-STn was associated with subsequent ovarian cancer (p=0.002). In the primary analysis restricted to cases within 5 years, the area-under-the-curve for CA-125-STn was 0.68; with moving window analysis suggesting retained predictiveness up to 5-7 years. Of the cases diagnosed within 5 years, 27% were in the top 5% of CA-125-STn levels (≥2.3 U/ml), with 35% and 49% of cases diagnosed in the top 10% and 20% respectively. Our findings were replicated by independent measurements of a separate aliquot from a subset of cases and controls. Neither CA-125, HE4 nor CA-125 glycovariants detected by MGL importantly predicted subsequent ovarian cancer.

**Conclusions and Relevance:** CA-125-STn can detect ovarian cancer several years before clinical diagnosis. Further research is warranted on larger cohorts with sequential samples to determine the shape of the relationship between CA-125-STn and risk, the value of longitudinal sampling of CA-125-STn, and whether other markers could be combined with CA-125-STn to more accurately predict ovarian cancer risk.

**Key Points:** *Question:* Can glycovariants of CA-125 detect ovarian cancer years before clinical diagnosis?

*Findings:* In a case-control study of 323 Swedish women followed longitudinally, the Sialyl-Thomsen-nouveau (STn) glycovariant of CA-125, but not conventional CA-125 or other markers, predicted ovarian cancer. Close to half of ovarian cancer cases within 5 years of blood sample occurred in the patients with the top 20% of CA-125-STn glycovariant levels.

*Meaning:* CA-125-STn can detect ovarian cancer several years before clinical diagnosis.

## Introduction

Randomized controlled trials have failed to find any benefit for CA-125 screening in reducing ovarian cancer mortality. In both the UKCTOCS^1^ and US PLCO^2^ trials, the lower bound of the 95% confidence intervals (CI) excluded any clinically relevant benefit for CA-125. One possible explanation for these findings is that changes in CA-125 concentration occur too late in the disease course to improve the outcome of treatment, such as by enabling surgical cure of early-stage disease. There was no difference in the stage distribution with screening in PLCO trial. In UKCTOCS, the reduction in stage III and IV disease was only 10%, too small to shift mortality even without the purported countervailing increased risk of mortality in early-stage patients in the screening arm^1^.

A second marker for ovarian cancer is human epididymal protein 4 (HE4)^3^. It can be combined with CA-125 to give a combined “risk of malignancy index”^4^. To date, there is a dearth of evidence that incorporating HE4 into ovarian cancer screening can lead to an important shift in stage at diagnosis.

It is known that glycan structures on CA-125 proteins are influenced by oncogenic processes^5^. Our hypothesis is that glycovariants of CA-125 would have greater specificity for ovarian cancer. In prior work, the glycovariant assays had better discrimination for ovarian cancer than the traditional CA-125 marker^6^. However, this study used the typical approach of assessing patients close to the time of ovarian cancer diagnosis, something which does not allow us to assess whether the markers can bring forward the date of diagnosis sufficiently to influence mortality. This is of particular relevance given recent findings the most common type ovarian cancer, high grade serous carcinoma, originates in the fallopian tubes^7,8^ and that by the time it is detected in the ovaries, it will likely have also spread intraperitoneally. Accordingly, early detection of cancer in the ovaries will already be too late to alter the course of disease.

Studies based on biorepositories containing cryopreserved samples from unscreened populations provide a better approach for evaluating prospective ovarian cancer markers, as the research blood sample is taken well before clinical diagnosis of cancer. We have previously used this approach to evaluate prostate specific antigen (PSA) and related kallikrein markers for prostate cancer using large prospective population-based observational cohorts in Malmö, Sweden. We were able to show that, for instance, while PSA has modest specificity for the short-term diagnosis of prostate cancer, it is extremely predictive of the long-term risk of prostate cancer death^9,10^.

Here we use samples and data from the same cohort to determine whether the CA-125 glycovariant assays, as well as conventional assays for CA-125 and HE4, could detect ovarian cancer several years before clinical diagnosis.

## Methods

### Study cohort

The prospective, population-based observational Malmö Diet and Cancer study (MDC) has been previously described^11^ and was approved by the Research Ethics Board at Lund University (LU 51-90 and 530-2008) with written informed consent obtained from each participant in accordance with the principles of the Declaration of Helsinki. In brief, 30,446 individuals (18,326 women) comprised the MDC baseline cohort, with 17,297 women giving blood between 1991-1996 at age 44 - 73 and having the minimum required 800 µl serum in the biobank. The MDC rescreening took place 2007-2012, with 2,168 women with sufficient serum, aged from 61 to 85 and giving blood an average of 17 years after baseline sample. Participants’ records were linked to the cancer registry at the National Board of Health and Welfare in Sweden to identify 82 women (74 at first sample, 8 at second sample) diagnosed with incident ovarian cancer up to 10 years after venipuncture. The registry has been shown to be highly accurate^12,13^. There were no early detection efforts for ovarian cancer for women in Malmö during the timeframe of this study.

Cases were matched 1:3 with controls randomly selected from MDC-participants who were within 6 months of age at blood draw, were sampled within 6 months of the index case and were alive and event free at the follow-up time at which the index case event occurred.

### Laboratory measurements

Pre-diagnostic cryopreserved aliquots of serum were thawed and measured using GLYVAR^®^ Ovarian I and II assays (Uniogen),and the conventional CA-125 (CanAg CA125 EIA, Fujirebio) and HE4 EIA (Fujirebio) assays according to the manufacturers’ instructions. The GLYVAR Ovarian I assay detects Sialyl-Thomsen-nouveau (STn) antigen; the GLYVAR Ovarian II assay detects STn, Tn (Thomsen-Nouveau) and disaccharide N,Nʹ-di-N-acetyllactose diamine (LacdiNAc) antigens by binding of Macrophage-Galactose-Lectin (MGL). Assays were conducted blind to case control status. CA125, HE4 and GLYVAR Ovarian I and II assays were conducted manually, using an automated immunoassay analyzer’s washer.

### Statistical approach

We used conditional logistic regression to assess the association between marker levels at baseline blood draw and subsequent diagnosis of ovarian cancer, including ICD-9 183.0 (malignant neoplasm of ovary), 183.2 (malignant neoplasm of fallopian tube, n=7) and 183.8 (malignant neoplasm of other specified sites of uterine adnexa, n=1), although the latter two were excluded in sensitivity analyses. In the primary analysis, we excluded patients who were diagnosed more than five years after blood draw, on the grounds that we would not expect those cases to be detectable by a blood marker. We also planned to exclude patients who had an aggressive diagnosis shortly after blood draw as these patient’s care would not be influenced by the marker values. However, only two cases died within two years after blood sampling and exclusion of those cases did not materially affect the results (see Supplementary table 1).

We also analyzed how time to diagnosis affected the association between markers and ovarian cancer by calculating the area-under-the-curve (AUC) when different cut-off levels were used for excluding cases based on the time between blood draw and diagnosis. We calculated the AUC for a moving two-year window of diagnosis centered between 1 and 9 years after blood draw, and then smoothed the results by linear regression with restricted cubic splines. To estimate the proportion of cases in a given population quantile of marker levels, we used the distribution of the marker values in cases as the numerator. We used the distribution of the marker levels in controls as the denominator because the incidence of disease is very low and because there was no association between marker levels and characteristics such as age. All markers were log transformed before analysis. Analyses were conducted using Stata 18 (Stata Corp., College Station, TX).

## Results

A total of 82 cases of incident ovarian cancer diagnosed within 10 years of blood draw were successfully matched with 3 controls. Three controls developed ovarian cancer after the follow-up time of the index case diagnosis, all of which occurred after 10 years. Samples could not be measured in one case and one control, leaving 81 cases, one of which had 2 controls. Table 1 shows characteristics of the cohort. Median age was 60, with about three-quarters of participants above the typical age of menopause. Most of the cases used the blood sample collected at enrollment in 1991 – 1996. Median levels of each marker were similar irrespective of case status; when restricted to cases diagnosed within 5 years from blood draw, there appears to be a difference only for CA-125-STn.

**Table 1.** Characteristics of the cohort. For the marker levels, only patients with the ICD-9 code for ovarian cancer and with evaluable marker levels are included (81 cases, 242 controls). Data are given as median (quartiles) or frequency (%).

| Characteristic | All participants |  |
| --- | --- | --- |
| Age at blood sample | 60 (52, 67) |  |
| Blood sample 2007 – 2011 | 29 (9.0%) |  |
|  | Cases | Controls |
| Time to diagnosis (years) | 4.9 (2.6, 7.5) |  |
| Primary site (ICD-9) |  |  |
| Ovarian (1830) | 74 (91%) |  |
| Fallopian tube (1832) | 6 (7.4%) |  |
| Other adnexal (1838) | 1 (1.2%) |  |
| CA-125 (U/mL) | 6.7 (4.8, 8.4) | 6.3 (4.3, 8.6) |
| HE4 (pmol/L) | 56.6 (45.9, 69.1) | 54.4 (44.1, 67.5) |
| CA-125-STn (U/mL) | 1.0 (0.7, 1.8) | 0.9 (0.6, 1.3) |
| CA-125-MGL (U/mL) | 0.3 (0.2, 0.6) | 0.3 (0.2, 0.5) |
| Where case diagnosed within 5 years (37 cases, 111 controls) |  |  |
| CA-125 (U/mL) | 6.9 (4.9, 8.5) | 6.0 (4.4, 8.5) |
| HE4 (pmol/L) | 51.7 (45.8, 68.8) | 54.2 (44.0, 65.9) |
| CA-125-STn (U/mL) | 1.3 (0.9, 2.2) | 0.9 (0.7, 1.3) |
| CA-125-MGL (U/mL) | 0.4 (0.2, 0.9) | 0.3 (0.2, 0.5) |

The association of marker levels with outcome is shown in table 2. The two established markers, CA-125 and HE4 showed no association with subsequent diagnosis of ovarian cancer either overall (up to ten years) or when cases diagnosed more than 5 years from blood draw were excluded. We also tried to restrict the cohort to cases diagnosed within a shorter time frame than 5 years but were unable to find any significant associations between levels of HE4 or the conventional CA-125 and subsequent diagnosis of ovarian cancer.

**Table 2.**
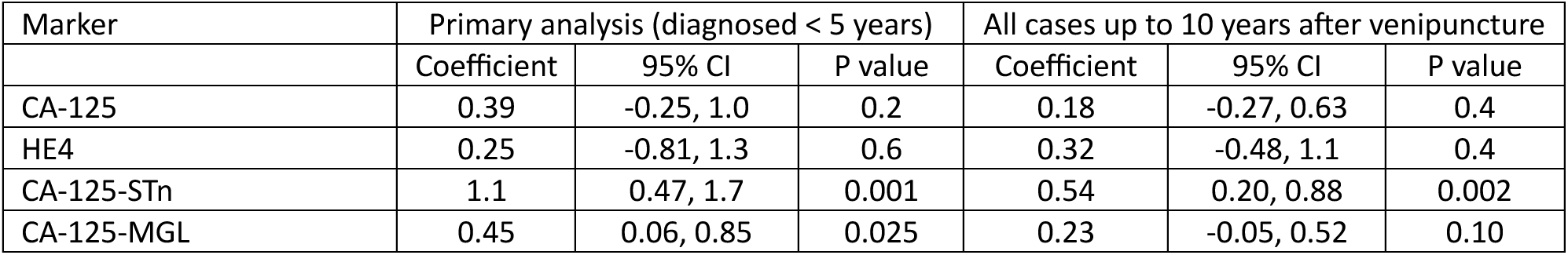
Univariable association of markers with case control status, by conditional logistic regression. Coefficients are given for 1 unit increase in the log of the marker (units of U/mL for CA-125 and glycovariants and pmol/L for HE4)

By contrast, serum levels of CA-125-STn are strongly associated with the risk of subsequent diagnosis of ovarian cancer within 5 years; statistical significance is maintained when including cases out to 10 years although the coefficient is smaller. Serum levels of CA-125-MGL binding sites are associated with subsequent diagnosis of ovarian cancer although the p-value does not remain significant in the context of multiple testing. Moreover, when we added CA-125-MGL binding sites to CA-125-STn in a multivariable model, it was no longer significant (p=0.7). This was perhaps not surprising given the correlation between the two glycovariants (ρ ∼0.4). Serum levels of CA-125-STn were essentially uncorrelated with CA-125 and HE4 (ρ < 0.15 for both) and these two markers did not add to CA-125-STn in a multivariable model (p>0.3 in all analyses).

The area-under-the-curve (AUC) for CA-125-STn for all cases diagnosed within 5 years from blood draw was 0.68. Figure 1 shows how the AUC varies over time using a moving window approach. The marker best discriminates early cases but maintains predictiveness up to 4 or 5 years out. Given the upper bound of the 95% CI, our results are consistent with CA-125-STnbeing able to predict cancers up to 7 or 8 years in advance of clinical diagnosis.

**Figure 1.**
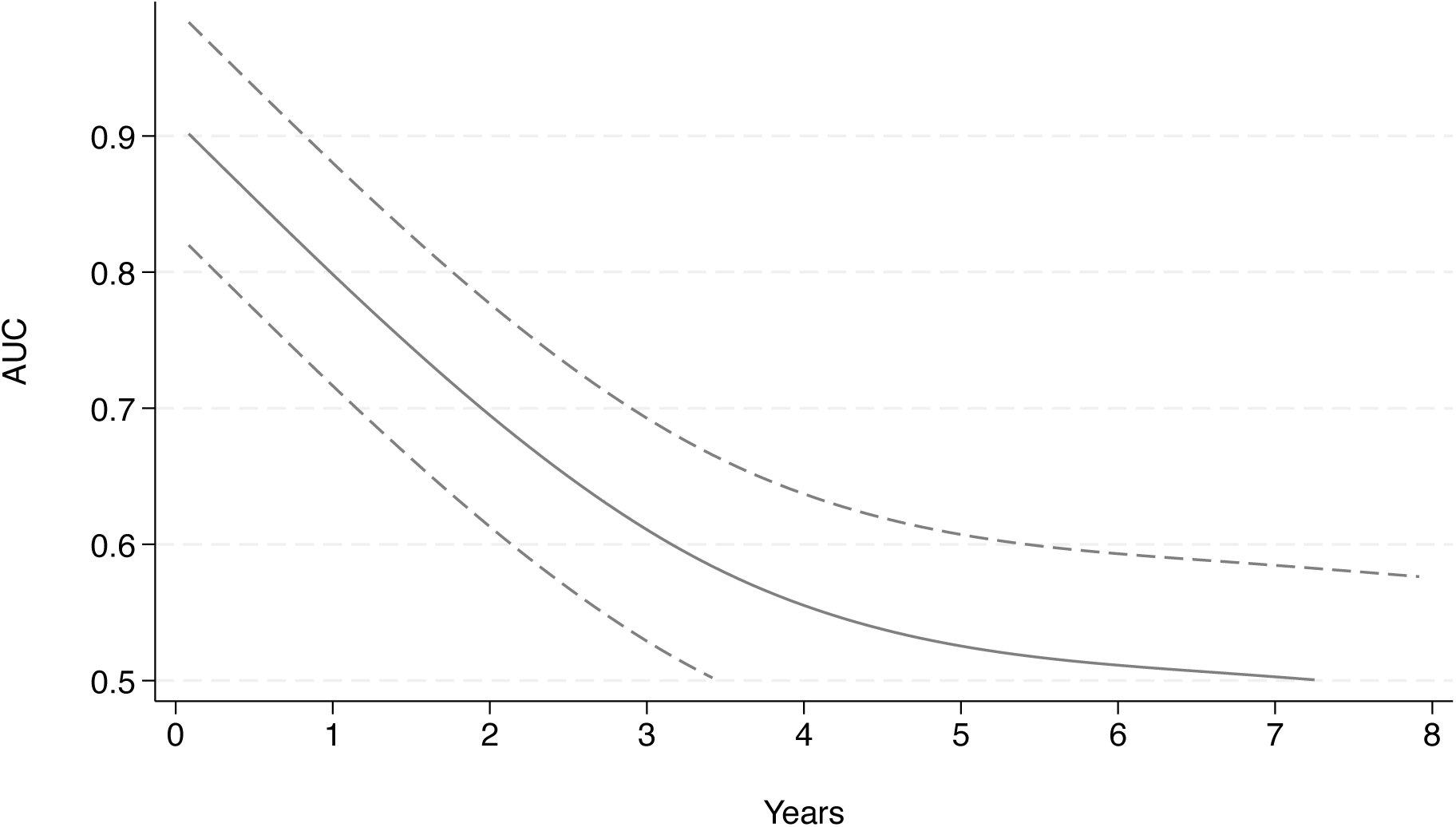
AUC of CA-125-STn for cases diagnosed at different times after blood sample. Analysis used a moving two-year window. For instance, the AUC at 4 years includes cases diagnosed 3 to 5 years after blood draw.

If the small number of fallopian tube and “other adnexal” cancers were excluded from the analysis, results were almost identical, with CA-125-STn statistically associated with outcome (p=0.001), no change in AUC (0.68) and no benefit to adding other markers.

Table 3 illustrates cut-points for CA-125-STn based on centiles of their distribution in controls, along with the proportion of incident ovarian cancer cases diagnosed within 5 years that had marker levels above each cut-point. Of the ovarian cancer cases diagnosed within 5 years, 27% had CA-125-STn levels in the top 5% (2.3 U/ml or higher) of the population. Women with a CA-125-STn concentration of 1.4 units/mL or higher in serum constitute 20% of the population but included nearly half of cases diagnosed with ovarian cancer in the subsequent five years.

**Table 3.**
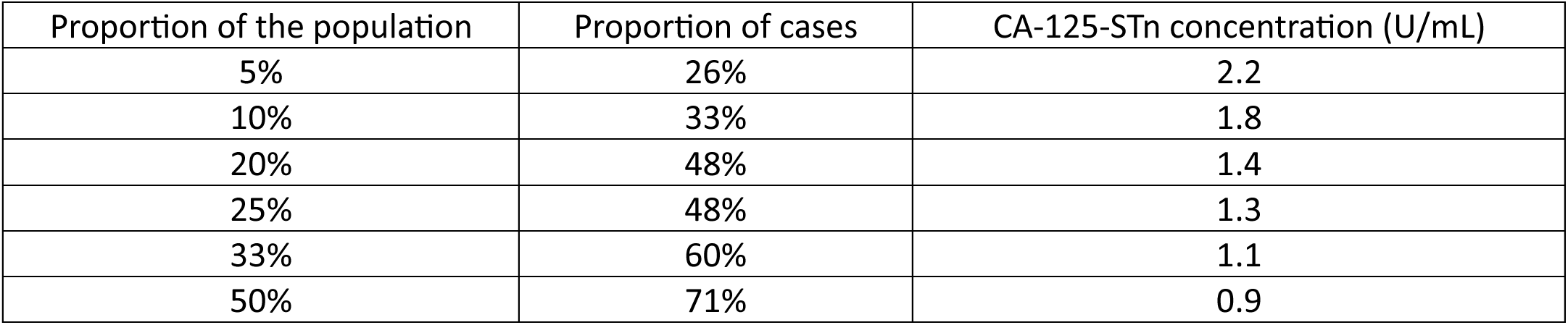
Cumulative proportion of cases at different population centiles of CA-125-STn.

| Proportion of the population | Proportion of cases | CA-125-STn concentration (U/mL) |
| --- | --- | --- |
| 5% | 26% | 2.2 |
| 10% | 33% | 1.8 |
| 20% | 48% | 1.4 |
| 25% | 48% | 1.3 |
| 33% | 60% | 1.1 |
| 50% | 71% | 0.9 |

Given the unprecedented nature of the findings, and some concerns about the levels observed in our initial analysis, possibly due to some technical issues with the washer of the automated instrument, we decided to remeasure both CA-125-STn and conventional CA-125 in the subset of our cohort with remaining sample sufficient for remeasurement. This second set of assays also used fresh assay materials for CA-125-STn. To avoid using up limited resources of cryopreserved samples unnecessarily, we restricted the analysis to cases diagnosed within 5 years of blood draw. Of the eligible 42 index cases and their matched controls, serum sample aliquots were available and analyzable for 35 cases and 87 controls. Results are shown in supplementary tables 2 and 3.

In brief, we replicated our original findings that the CA-125-STn predicted subsequent diagnosis of ovarian cancer. We also found that CA-125 was predictive, although this was driven by three cases of ovarian cancer diagnosed shortly after blood draw. The three highest CA-125 levels were 62 U/ml, 37 U/ml, and 34 U/ml, with ovarian cancer diagnosed after 13 months, <1 month and 7 months from blood draw, respectively. The AUC for CA-125 - but not for CA-125-STn levels - was markedly reduced by the exclusion of cases diagnosed within 1 or 2 years after blood draw (0.62, 0.59 and 0.56 for CA-125 including all cases, and excluding those after 1 and 2 years respectively). Corresponding AUCs for CA-125-STn levels were 0.69 including all cases, 0.72 excluding cases diagnosed within one year from blood draw, and 0.74 excluding cases diagnosed within two years from blood draw. If cases diagnosed within two years were excluded, only CA-125-STn (p= 0.002) but not CA-125 (p=0.2) retained predictiveness. We also found evidence that the discriminatory accuracy of the CA-125-STn assay was superior to that of the original measurements (AUC of 0.69 vs. 0.65 including only samples that were independently remeasured). As the number of cases and controls was limited, we were unable to conduct the analyses looking at the cumulative distribution of cases by population quantile of CA-125-STn nor use the “moving window” approach to assess the timeframe of predictiveness.

## Discussion

We found that serum levels of a glycovariant of CA-125 can predict risk of ovarian cancer diagnosis up to about 5 years before clinical diagnosis in women not subject to any form of ovarian cancer screening. This result was replicated by an independent remeasuring of a subset of cryopreserved serum samples. About half of the ovarian cancers diagnosed within 5 years occurred in women in the top 20% of CA-125-STn levels. Neither conventional CA-125, HE4 or CA125-MGL binding sites measured in serum were able to offer clinically relevant prediction of subsequent ovarian cancer.

Prior research on CA-125 and HE4 has looked predominately at prevalent rather than incident cancer. In what is generally regarded as the first major study on CA-125 in 1983^14^, blood levels of patients with “surgically demonstrated ovarian carcinoma”, ∼85% of whom had stage III or IV disease were compared with male and female healthy controls, and male and female patients with a variety of non-malignant diseases. The cut-off still widely used in contemporary practice - 35 U/mL - was derived from this original study. Well-known studies on HE4 similarly compared healthy controls to women known to have ovarian cancer^15,16^.

Recent developments suggest that high-grade serous ovarian carcinoma actually originates in the fallopian tubes.^7,8^ For instance, studies of fallopian tubes removed during salpingo-oophrectomy demonstrate a series of stepwise changes in fallopian tissue, from p53 loss, to serous tubal intraepithelial carcinoma (STIC), to carcinoma invading the basement membrane, and on to metastasis. Estimates of the timeframe for progression of STIC to ovarian cancer are about 6 years^8,17^. Further confirmatory evidence for a fallopian tube origin for ovarian cancer is that women without fallopian tubes are at much reduced risk: a recent systematic review concluded reported an 80% risk reduction”^18^. This suggests an obvious clinical strategy of testing women for CA-125-STn and encouraging salpingectomy, a safe and benign procedure^19^, for those with elevated levels.

Power limitations meant that we were unable to determine whether the relative proportion of the different markers – such as in the free-to-total PSA ratio – might be of value. Moreover, given that we only had access to a single blood sample, we were also unable to use the Risk of Ovarian Cancer Algorithm (ROCA), the method used in the UKCTOCS and other trials, which depends on sequential CA-125 samples^20^. It seems likely that ROCA would have had superior performance to a single CA-125 value, but by the same token, the same might be true of CA-125-STn. Further research should examine longitudinal changes in CA-125-STn in sequential samples: it seems reasonable to suppose that a longitudinal algorithm for CA-125-STn would be superior to one for CA-125 given that a single value of the former, but not the latter, is predictive.

A final limitation of our study is that we do not have patient-level data on histology or stage, although given the age distribution, most of the cases are likely to be high-grade serous carcinoma^20^. Accordingly, we are unable to determine whether the patients identified by the CA-125-STn assay would have experienced an improved chance of cure from early detection with a biomarker test. It could be that CA-125-STn increases before a tumor could be detected on further work-up or, alternatively, only after cancer has spread intraperitoneally. Further research is required to elucidate the role of CA-125-STn in early detection of ovarian cancer by looking at stage and histology at diagnosis. Further research should also examine the value of CA-125-STn when combined with ROCA or even a multi-cancer early detection test (MCED)^21^.

In sum, we have provided the first evidence that a blood marker, CA-125-STn, can predict ovarian cancer several years before clinical diagnosis. Further research is warranted on larger cohorts with sequential samples to determine the shape of the relationship between CA-125-STn and risk, the value of longitudinal sampling of CA-125-STn, and whether other markers could be combined with CA-125-STn to estimate ovarian cancer risk more accurately.

## Data Availability

All data produced in the present study are available upon reasonable request to the authors
All data produced in the present work are contained in the manuscript

---

**Supplementary table 1.** Univariable association of markers with case control status, by conditional logistic regression.

| Marker | Primary analysis (diagnosed < 5 years) |  |  | Include only early cases (diagnosed <5 years but no cancer specific death < 2 years) |  |  |
| --- | --- | --- | --- | --- | --- | --- |
|  | Coefficient | 95% CI | P value | Coefficient | 95% CI | P value |
| CA-125 | 0.39 | -0.25, 1.04 | 0.2 | 0.42 | -0.23, 1.07 | 0.2 |
| HE4 | 0.25 | -0.81, 1.31 | 0.6 | 0.38 | -0.73, 1.49 | 0.5 |
| CA-125-STn | 1.09 | 0.47, 1.70 | 0.001 | 1.07 | 0.45, 1.69 | 0.001 |
| CA-125-MGL | 0.45 | 0.06, 0.85 | 0.025 | 0.47 | 0.06, 0.88 | 0.024 |

**Supplementary table 2.**
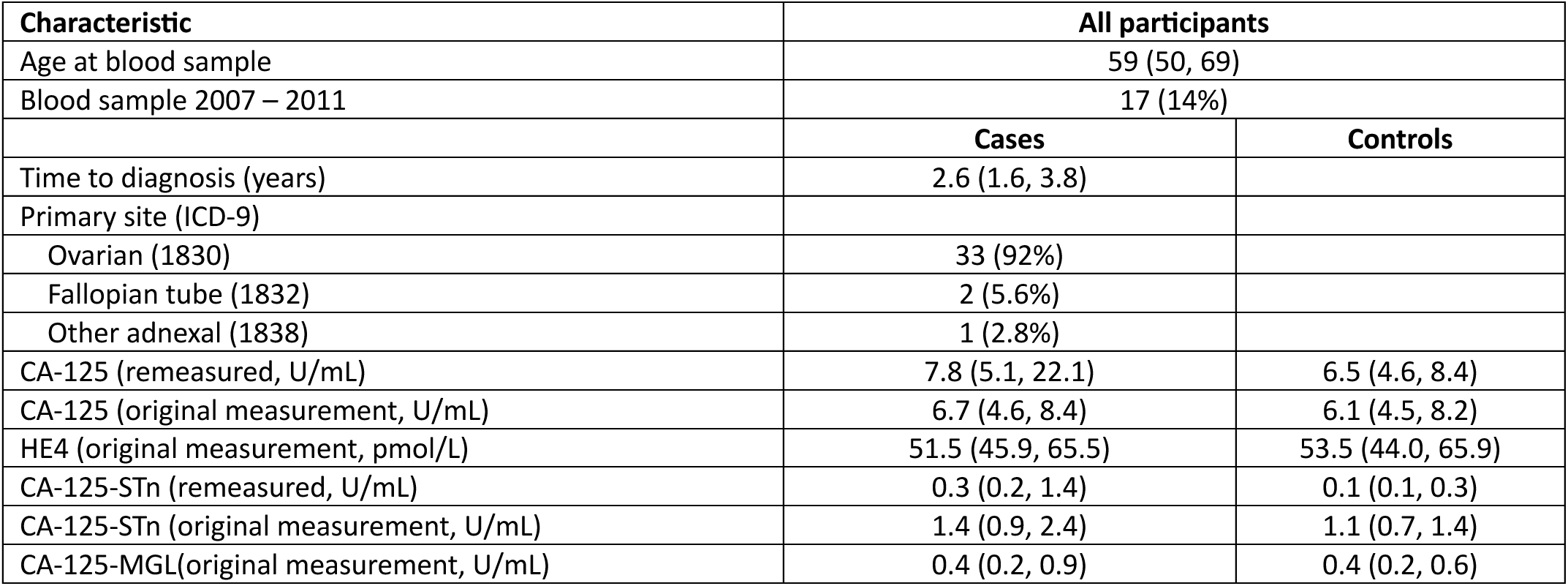
Characteristics of the cohort. There were 36 cases and 89 controls (3, 13 and 20 cases with 1, 2 and 3 controls respectively). Markers levels are given for cases irrespective of ICD-9 code. Data are given as median (quartiles) or frequency (%).

**Supplementary Table 3.** Univariable association of markers with case control status, by conditional logistic regression. Coefficients are given for 1 unit increase in the log of the marker (units of U/mL for CA-125 and glycovariants and pmol/L for HE4)

| Marker | Primary analysis (diagnosed < 5 years) |  |  |
| --- | --- | --- | --- |
|  | Coefficient | 95% CI | P value |
| CA-125 (remeasured) | 0.99 | 0.31, 1.66 | 0.004 |
| CA-125 (original measurement) | 0.00 | -0.07, 0.08 | 1 |
| HE4 | 0.09 | -1.08, 1.26 | 0.9 |
| CA-125-STn (remeasured) | 0.59 | 0.24, 0.93 | 0.001 |
| CA-125-STn (original measurement) | 0.58 | 0.03, 1.13 | 0.040 |
| CA-125-MGL | 0.48 | 0.03, 0.93 | 0.038 |

